# Multimodal Deep Learning Framework for Customizable and Interpretable Parkinson’s Disease Detection

**DOI:** 10.64898/2026.01.01.25343252

**Authors:** Mitesh Kothari, Ganeshan Arumuganainar, Kaliraj Konar

## Abstract

**Background:** Parkinson’s Disease (PD) is often reduced to its most visible motor symptoms, yet it is a systemic neurodegenerative disorder with a highly heterogeneous presentation. While cardinal motor signs such as bradykinesia, rigidity, and tremor arise from the loss of dopaminergic neurons in the substantia nigra, they typically manifest only after substantial neurodegeneration (approximately 50–70% loss) has already occurred, inevitably leading to delayed detection [1] PD significantly impacts non-motor and fine-motor domains as well that are frequently overlooked. Research indicates that hypokinetic dysarthria (voice impairment) affects approximately **89%** of PD patients, often as an early prodromal sign [18]. Similarly, micrographia (handwriting impairment) is observed in up to **63%** of cases, while non-motor symptoms such as hyposmia (loss of smell) and REM sleep behavior disorder occur in over **70%** and **40%** of patients, respectively—often years before clinical diagnosis [19, 20]. Consequently, diagnostic systems that rely on a single modality fail to capture this complexity, leading to missed detections in patients whose primary symptoms fall outside that specific domain. To address this, we propose a holistic, multimodal AI framework that explicitly targets these diverse pathological vectors—Voice, Gait, Handwriting, and Non-Motor Symptoms—to ensure robust and early detection across the full spectrum of the disease.

**Methods:** We propose a modular multimodal AI framework that integrates five complementary inputs: voice recordings, signals captured with the help of a smart pen during drawing spiral/meander, hand-drawn spiral/meander images, wearable sensor-driven gait data, and MDS-UPDRS questionnaire-derived symptom scores. Each modality undergoes an independent preprocessing and specialized modeling pipeline. Outputs from these specialized models are combined using a weighted aggregation engine, which allows for customizable contribution of each modality to the final classification.

**Results:** Preliminary experiments show that the unimodal pipelines achieved high accuracy, with the Random Forest (Voice) achieving 89%, XGBoost (Drawing Signal) up to 93%, and ResNet-18 (Drawing Image) up to 92%. Incorporating the Transformer model for gait data, which achieved 86% accuracy, significantly boosts the detection of subtle motor deficits. The proposed approach is expected to improve the overall diagnostic sensitivity and specificity relative to any unimodal baseline, offering transparent score breakdowns for clinical use.

**Conclusion:** This study validates a comprehensive, multimodal Machine Learning framework designed to capture the holistic nature of clinical Parkinson’s Disease. Our results indicate that fine motor control—analyzed through both dynamic handwriting signals and static imagery—serves as a highly discriminative biomarker, offering superior detection of subtle kinematic tremors. Furthermore, the integration of vocal analysis and spatiotemporal gait modeling ensures that the system captures the full spectrum of pathology, distinguishing between phonatory deficits and gross motor irregularities. By synthesizing these diverse clinical indicators, the proposed architecture overcomes the sensitivity limitations of single-modality systems, establishing a robust, non-invasive foundation for objective early screening and longitudinal patient monitoring in real-world settings.

## 1 Introduction

### 1.1 Context and Background

Parkinson’s Disease (PD) is the second most common neurodegenerative disorder, primarily characterized by motor symptoms such as bradykinesia, rigidity, tremor, and postural instability, arising from the loss of dopaminergic neurons in the substantia nigra. The impact extends beyond motor functions, encompassing debilitating non-motor symptoms (NMS) like hyposmia, sleep disorders, and mood disturbances, which often predate motor onset by several years [6, 2].

The application of Artificial Intelligence (AI) and Machine Learning (ML) in healthcare has opened promising avenues for automated diagnostics, particularly in analyzing complex, high-dimensional data. In the context of PD, this is critical because conventional clinical diagnosis, while essential, relies heavily on subjective assessments (like the Movement Disorder Society-Unified Parkinson’s Disease Rating Scale, MDS-UPDRS), which can be variable, time-consuming, and prone to diagnostic delays, especially in the early stages [3].

### 1.2 Problem Statement

Current diagnostic protocols rely heavily on subjective clinical assessments, such as the MDS-UPDRS, which require observation by specialized neurologists. These traditional methods are inherently reactive; a diagnosis is typically confirmed only after the manifestation of cardinal motor signs, by which point approximately 50–70% of nigral dopaminergic neurons have already been lost [1]. Furthermore, this dependence on specialized in-person examinations creates significant barriers in resource-limited settings, leading to diagnostic delays and high rates of misclassification in the early stages of the disease [3].

While digital biomarkers offer a promising alternative, existing machine learning approaches suffer from critical deficiencies. Most current models are *unimodal*, analyzing isolated data streams (e.g., only voice or only gait), which fails to capture the heterogeneous and systemic nature of Parkinson’s pathology. Additionally, high-performing Deep Learning models often operate as “black boxes,” lacking the interpretability required for clinical adoption. To address these gaps, this study proposes an open-source, *multimodal* framework that integrates voice, gait, handwriting, and non-motor symptoms. By synthesizing these diverse inputs into an interpretable diagnostic score, the framework aims to democratize PD screening and reduce diagnostic delay without reliance on expensive commercial hardware.

### 1.3 Knowledge Gap

Current voice-based and multimodal PD detection systems suffer from significant limitations. Most existing approaches rely on single-modality analysis (voice or gait or drawing in isolation), missing complementary biomarkers that enhance diagnostic confidence [21]. Existing machine learning models lack interpretability—clinicians cannot understand which acoustic or motor features drove a specific diagnosis.

### 1.4 Objective and Significance

The objective of this work is to propose and implement a modular, multimodal AI framework for the automated risk stratification of Parkinson’s Disease. This framework processes five distinct data modalities—voice, hand-drawing signals, handwriting images, gait, and non-motor symptoms—using specialized ML/Deep Learning pipelines.

The significance of this solution lies in its design for **customization and interpretability**. By utilizing a strategy with adjustable weights it can provide a transparent breakdown of the contribution of each digital biomarker, moving the field towards a scalable and ethical AI solution for neurodegenerative disease management.

## 2 Methods

### 2.1 General Machine Learning Workflow

Irrespective of the specific data modality employed, our research methodology adheres to a standard, iterative Machine Learning project lifecycle. This process ensures rigor and reproducibility across all subsystems of the framework:

**Figure 1:**
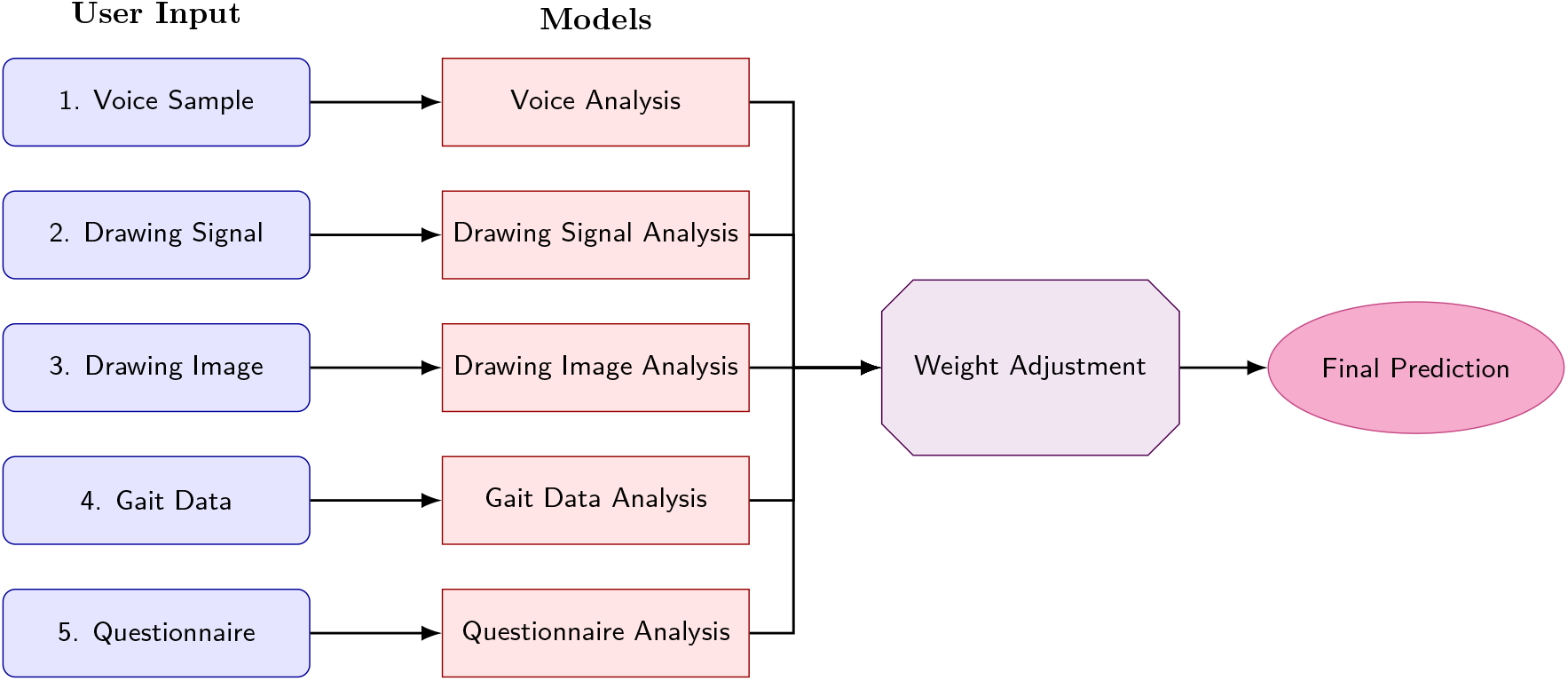
Architecture of the Proposed Multimodal Framework. Five distinct modalities are processed by specialized sub-models, and their outputs are aggregated via a weighted voting mechanism to produce the final diagnostic prediction.

1. **Problem Definition:** The cycle begins with defining the specific Business Problem and framing it as a Machine Learning Task (e.g., binary classification of healthy vs. pathological samples).
2. **Data Pipeline:** We proceed to Data Collection and Preparation, ensuring high-quality inputs. This phase includes a feedback loop for Data Augmentation to address class imbalances or data scarcity.
3. **Feature Engineering:** Raw data is transformed into informative representations. This stage is iterative; if model performance is insufficient, we loop back to perform Feature Augmentation or refine the selection logic.
4. **Modeling:** The core phase involves Model Training and Parameter Tuning (hyperparameter optimization) to minimize loss functions specific to the architecture (e.g., Cross-Entropy, LogLoss).
5. **Evaluation and Deployment:** Models undergo rigorous Evaluation against hold-out test sets. If the business goals (accuracy, sensitivity, specificity targets) are met, the model proceeds to Testing and Deployment. If not, the workflow reverts to the Feature Engineering or Data Collection phases for refinement.
6. **Monitoring:** Post-deployment, a Monitoring and Debugging loop tracks real-world performance, triggering retraining with new data when necessary to prevent concept drift.

### 2.2 Multimodal Aggregation and Weight Adjustment

#### 2.2.1 Traditional Fusion Approaches

Traditional multimodal machine learning systems typically employ one of two fusion strategies: *Early Fusion* (feature concatenation) or *Late Fusion* (static ensemble voting). In Early Fusion, raw features from all modalities are merged into a single large vector before classification. While this captures interactions between modalities, it suffers from the “curse of dimensionality” and requires all sensors to be present simultaneously [**?**]. In standard Late Fusion, the decisions from independent models are combined using fixed rules, such as Majority Voting or static averaging. These traditional methods are rigid; they treat all sensors as equally reliable at all times, failing to account for scenarios where a specific sensor may be noisy, missing, or clinically irrelevant (e.g., a patient with a non-PD leg injury providing unreliable gait data) [**?**].

#### 2.2.2 Proposed Dynamic Weighted Aggregation

To address the lack of flexibility in traditional fusion, we implement a **Dynamic Weighted Aggregation** engine. Unlike static voting, our framework assigns a confidence weight (*w*_*i*_) to the probability output (*P*_*i*_) of each modality model *i*. The final risk score *S*_*final*_ is calculated as:

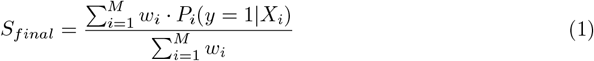

This architecture introduces two novel capabilities:

a. **Human-in-the-Loop Customization:** Clinicians can manually adjust weights based on the availability or quality of data. For example, if a patient cannot perform the gait task due to arthritis, the physician can set *w*_*gait*_ = 0, normalizing the prediction based solely on voice and handwriting.
b. **Interpretability:** By explicitly exposing the contribution of each modality to the final score, the system moves away from “black-box” diagnosis toward “explainable AI,” a critical requirement for medical ethics and trust [**?**].

This structured approach is applied individually to the Voice, Handwriting, Gait, and Questionnaire modules detailed below.

### 2.3 Voice Recording Analysis

#### 2.3.1 Dataset

The **Mobile Device Voice Recordings at King’s College London (MDVR-KCL)** dataset is utilized for voice analysis. The dataset comprises recordings from 37 participants, divided into 21 Healthy Controls (HC) and 16 Parkinson’s Disease (PD) patients [8].

We selected the MDVR-KCL dataset specifically for its high ecological validity in the context of telemedicine and remote patient monitoring. Unlike datasets collected in soundproof studio booths, MDVR-KCL captures voice data using consumer-grade hardware (smartphones) held in close proximity to the ear. This setup mimics real-world conditions, ensuring that our detection model is robust enough for deployment in accessible, low-resource mobile health applications.

From these raw acoustic waveforms, we capture distinct vocal biomarkers indicative of PD-induced dysphonia and hypophonia. The feature extraction process targets two primary categories of acoustic characteristics:

- **Phonation and Perturbation Features:** We extract metrics such as *Jitter* (micro-fluctuations in pitch period) and *Shimmer* (micro-fluctuations in amplitude). These features quantify the instability of vocal fold vibration, a common early symptom of Parkinsonian speech caused by reduced motor control over the laryngeal muscles.
- **Spectral and Cepstral Features:** We compute *Mel-Frequency Cepstral Coefficients (MFCCs)* and *Harmonics-to-Noise Ratio (HNR)*. These features capture the spectral envelope and the ratio of periodic to aperiodic components in the voice, effectively characterizing the “breathiness” and articulatory degradation often observed in PD patients.

Data collection occurred between September 26 and 29, 2017, at King’s College London Hospital. Recordings were conducted in a standardized examination room (approx. 10 m^2^ with 500 ms reverberation time) using a Motorola Moto G4 Smartphone via the “Toggle Recording App.” The resulting audio files are saved in WAVE format (.wav) with high-fidelity specifications: 44.1 kHz sample rate and 16-bit depth.

#### 2.3.2 Voice Data Preprocessing

The preprocessing workflow transforms raw audio into tabular features using the following pipeline:

1. **Data Organization and Loading** Audio files are structured by speech type (phonation vs. reading) and diagnostic group (HC vs. PD). Utilizing the `librosa` python library [14], files are loaded with a standardized duration cap of 30 seconds to ensure consistency across the cohort.
2. **Acoustic Feature Extraction** We extract four categories of features to characterize vocal impairment:
  - **Jitter Metrics (Frequency Perturbation):** To quantify cycle-to-cycle variation in the fundamental frequency (*F*_0_), pitch contours are extracted using `librosa.piptrack`. Metrics include Mean *F*_0_, *F*_*hi*_, *F*_*lo*_, Percent Jitter, Absolute Jitter, Relative Average Perturbation (RAP), Pitch Perturbation Quotient (PPQ), and Degree of Voice Breaks (DDP).
  - **Shimmer Metrics (Amplitude Perturbation):** The Short-Time Fourier Transform (STFT) is computed to extract amplitude envelopes. Metrics include Shimmer (dB), Amplitude Perturbation Quotient (APQ3, APQ5, Overall APQ), and Difference of Differences of Amplitudes (DDA).
  - **Noise Metrics:** To characterize breathiness, we compute the Noise-to-Harmonics Ratio (NHR) and Harmonics-to-Noise Ratio (HNR) via spectral peak detection.
  - **Nonlinear Complexity:** To capture signal dynamics, we calculate Recurrence Period Density Entropy (RPDE), Detrended Fluctuation Analysis (DFA), Correlation Dimension (*D*_2_), and Pitch Period Entropy (PPE).
3. **Output Generation** Features are aggregated into a `pandas.DataFrame`, labeled (HC=0, PD=1), and cleaned of malformed entries before being exported to CSV for machine learning ingestion.

#### 2.3.3 Model Development

Following feature extraction, a supervised machine learning pipeline was constructed to classify subjects into Healthy Controls (HC) and Parkinson’s Disease (PD) groups. The modeling process was implemented using the Scikit-learn library in Python.

1. **Data Partitioning** To ensure the model’s generalizability and prevent data leakage, the dataset was subjected to a stratified train-test split.
  - **Input Preparation:** Identifiers were removed from the dataset, isolating the acoustic features (*X*) and the diagnostic labels (*y*).
  - **Stratified Splitting:** The data was partitioned into a training set (80%) and a testing set (20%) using a fixed random seed (`random_state=42`) for reproducibility. Crucially, stratified sampling (`stratify=y`) was employed to ensure that the ratio of PD patients to Healthy Controls remained consistent across both training and testing subsets, preventing bias in the model evaluation.
2. **Classifier Configuration** A Random Forest Classifier (RFC) was selected as the inference engine due to its ensemble nature, which reduces the risk of overfitting common in small medical datasets, and its ability to handle non-linear relationships between acoustic features. The model was instantiated with the following hyperparameters:
  - **Ensemble Size (**`n_estimators`): 100 decision trees were utilized to ensure stability in predictions.
  - **Tree Depth (**`max_depth`): Nodes were allowed to expand until all leaves were pure (`None`), capturing maximum detail from the training data.
  - **Splitting Criteria:** The minimum number of samples required to split an internal node was set to 2 (`min_samples_split=2`), with a minimum of 1 sample required at a leaf node (`min_samples_leaf=1`).
3. **Evaluation and Interpretability Framework** The model performance was evaluated on the unseen testing set (*X*_*test*_) using a multi-faceted approach:
  - **Performance Metrics:** The classification efficacy was measured using standard metrics generated via a classification report, including Precision, Recall, F1-Score, and overall Accuracy.
  - **Error Analysis:** A Confusion Matrix was generated to visualize the distribution of True Positives, True Negatives, False Positives, and False Negatives, allowing for an analysis of the model’s sensitivity in detecting PD.
  - **Feature Importance:** To identify the most significant acoustic biomarkers, Gini impurity-based feature importance scores were extracted from the trained ensemble. The top 10 most influential features were ranked and visualized to provide clinical interpretability to the blackbox model.

Finally, the trained model, feature metadata, and performance metrics were serialized and saved to facilitate future deployment and inference consistency.

#### 2.3.4 Results

The Random Forest model achieved an overall accuracy of 89% on the test set. The classification report revealed a precision of 0.89 and a recall of 0.89 for the PD class, indicating the model’s ability to correctly identify pathological samples.

### 2.4 Hand-Drawing Signal Analysis

#### 2.4.1 Dataset Description

For the kinematic analysis modality, we utilize the **NewHandPD** dataset [9], an enhanced iteration of the original HandPD corpus. This dataset comprises distinct recording sessions from 66 individuals, stratified as follows:

- **Healthy Control (HC) Group:** 35 individuals (18 male, 17 female) with an average age of 44.05 ± 14.88 years.
- **Patient (PD) Group:** 31 individuals (21 male, 10 female) with an average age of 57.83 ± 7.85 years.

Data acquisition was performed using a Biometric Smart Pen (BiSP). While the original data collection protocol included various shapes (such as circles), this study focuses exclusively on the **Spiral** and **Meander** exams due to their high data completeness and clinical relevance.

The NewHandPD dataset is selected for this research because it provides access to *dynamic* timeseries data, rather than relying solely on static offline images. By utilizing the Biometric Smart Pen sensors, the dataset captures temporal information—such as pen pressure, velocity, and in-air movement—which are lost in standard image scanning. This is crucial for detecting *bradykinesia* (slowness of movement) and *micrographia* (progressive reduction in writing amplitude), which are hallmark motor symptoms of Parkinson’s Disease.

From these multivariate time-series signals recorded at 1000 Hz, we extract features targeting two specific physiological phenomena:

- **Tremor Detection (Frequency Domain):** We analyze the 4–7 Hz frequency band to identify the specific “resting tremor” characteristic of PD.
- **Motor Irregularity (Time Domain):** We calculate metrics related to signal energy and zero-crossing to quantify the fine motor jitter and lack of smoothness in the patient’s hand movements.

#### 2.4.2 Signal Preprocessing and Feature Extraction

The pipeline separates Spiral and Meander patterns and extracts the following features in both frequency and time domains.

1. **Frequency-Domain Features (Tremor Analysis)** Parkinsonian tremor typically manifests in the 4–7 Hz range. We apply the Fast Fourier Transform (FFT) to the signal *x*(*n*). Let *X*(*f*) be the frequency spectrum obtained via FFT.
  - **Dominant Frequency (***F*_*dom*_): The frequency associated with the maximum spectral energy.

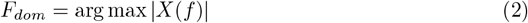
  - **Tremor Band Power (***P*_*tremor*_): The mean spectral magnitude within the physiological tremor band (4–7 Hz).

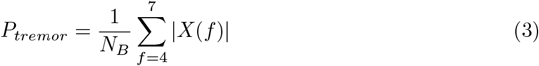

where *N*_*B*_ is the number of frequency bins in the range.
2. **Time-Domain Features** To capture motion irregularity, the following metrics are calculated:
  - **Zero-Crossing Rate (ZCR):** Measures the frequency of sign changes, reflecting fine motor jitter.

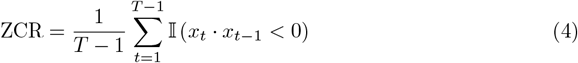

where 𝕀 is the indicator function and *T* is the total time steps.
  - **Signal Energy (***E*_*s*_): Quantifies movement intensity and force.

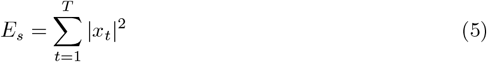
  - **Peak Count (***N*_*p*_): Measures tremulous activity by counting local maxima (peaks) that exceed a specific prominence threshold *δ*.

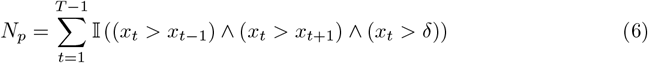

#### 2.4.3 Feature Selection and Optimization

Following the extraction of metrics, a rigorous dimensionality reduction process was applied to the tabular data to prevent the “curse of dimensionality” and improve model inference speed.

- **Variance Filtering:** Features exhibiting zero variance (constant values across all samples) were identified and removed.
- **Correlation Thresholding** A Pearson correlation analysis was conducted between each feature and the diagnostic target. To ensure only highly discriminative predictors were retained, features with an absolute correlation coefficient |*r*| *<* 0.35 were discarded. This step significantly reduced noise, isolating specific kinematic markers strongly associated with Parkinsonian symptoms.

#### 2.4.4 Algorithmic Framework: XGBoost

Based on comparative experimentation with Support Vector Machines (SVM), Logistic Regression, and Random Forests, the **eXtreme Gradient Boosting (XGBoost)** algorithm [11] was selected as the optimal classifier for this modality.

XGBoost was chosen for its gradient-boosted decision tree architecture, which effectively models non-linear interactions between kinematic features while robustly handling potential outliers in the signal data. The model was trained using the Logarithmic Loss (`logloss`) objective function to optimize class probability estimates.

#### 2.4.5 Training and Validation Protocol

The optimized dataset was partitioned into training (70%) and testing (30%) sets. Stratified sampling was employed to maintain the class distribution balance between Healthy Controls and PD patients. For the inference pipeline, the specific feature subset identified during the training phase was serialized (saved as `.npy` files) to ensure that incoming test data undergoes the exact same feature filtering process as the training data.

#### 2.4.6 Results: Signal Analysis

The XGBoost classifier was evaluated separately on signals derived from Spiral drawings and Meander drawings.

- **Spiral Signal Performance:** The dynamic analysis of Spiral drawings yielded an accuracy of **91%**, with a Precision of **92%** and Recall of **89%**.
- **Meander Signal Performance:** The Meander drawing signals resulted in an accuracy of **94%**, with a Precision of **95%** and Recall of **92%**.

### 2.5 Hand Drawing Image Analysis

#### 2.5.1 Dataset Description

For the static visual analysis of handwriting, we utilize the **NewHandPD** dataset [9], an extended and refined version of the original HandPD corpus. The dataset was collected at the Botucatu Medical School (São Paulo State University, Brazil) and comprises data from **66 participants**, stratified into two groups:

- **Healthy Control (HC) Group:** 35 individuals (18 male, 17 female; average age 44.05 ± 14.88 years).
- **Patient (PD) Group:** 31 individuals (21 male, 10 female; average age 57.83 ± 7.85 years).

Each participant performed specific motor tasks using a Biometric Smart Pen (BiSP), including drawing **Archimedean Spirals** and **Meanders** (complex looping patterns). The resulting drawings were digitized into high-resolution binary images, providing a static visual record of the movement trajectory.

We selected the NewHandPD dataset because it specifically targets the visual manifestations of *bradykinesia* and *tremor* in a static format, mimicking traditional pen-and-paper clinical screening. Unlike dynamic signals which require specialized sensors, static images can be analyzed using standard computer vision techniques, increasing the accessibility of the screening tool.

The dataset allows our Deep Learning models to capture two critical morphological features:

- **Micrographia:** A progressive reduction in handwriting size and amplitude, a hallmark early symptom of PD.
- **Tremor Artifacts:** Involuntary oscillations that appear as “jagged” or “shaky” lines in the static trace, distinguishable from the smooth curves produced by healthy controls.

#### 2.5.2 Image Preprocessing Pipeline

The raw image dataset, divided into Spiral and Meander subsets, underwent a standardized preprocessing pipeline utilizing the `torchvision` library.

- **Resizing:** All input images were resized to spatial dimensions of 224 × 224 pixels to align with the input requirements of the ResNet architecture.
- **Normalization:** Tensor values were normalized using the standard ImageNet statistics (Mean, Std) to facilitate faster convergence during gradient descent.
- **Augmentation Policy:** Consistent with the specific nature of medical handwriting analysis, geometric augmentations (such as random rotations or flips) were explicitly excluded. This ensures that the directional integrity of the patient’s tremors and stroke patterns remains unaltered.

**Figure 2:**
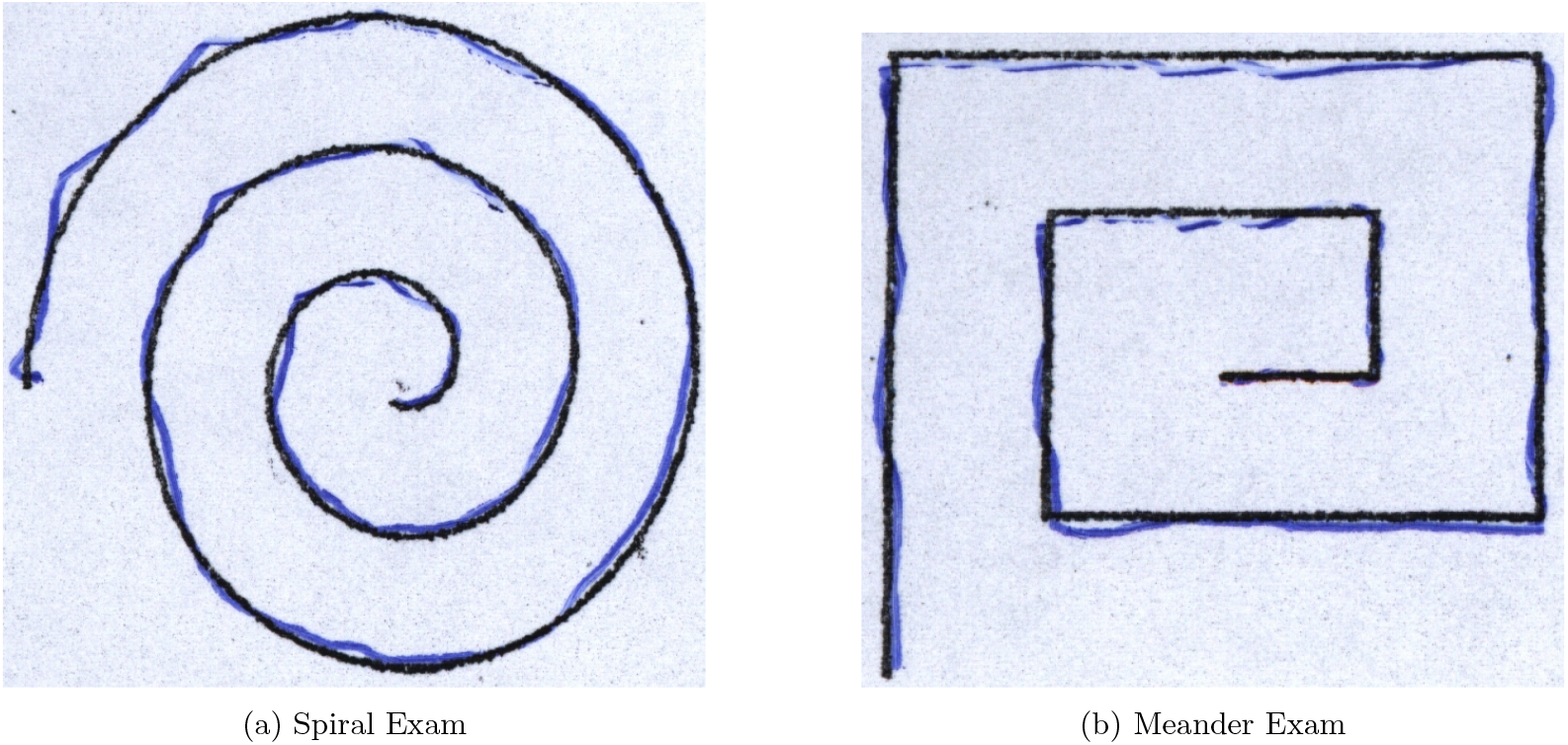
Sample preprocessed images from the NewHandPD dataset [9]. (a) An Archimedean spiral exhibiting smooth curvature. (b) A Meander pattern used to test complex cornering and fine motor control. Both of the samples are from a healthy individual.

#### 2.5.3 Convolutional Neural Network Architecture

A Transfer Learning approach was adopted to leverage feature extractors learned from large-scale datasets. We utilized the **ResNet-18** architecture, a deep Residual Network pre-trained on the ImageNet dataset.

#### 2.5.4 Model Configuration

The architecture was modified for binary classification (Parkinson’s vs. Non-Parkinson’s) as follows:

a. **Feature Extractor:** The convolutional layers of ResNet-18 were retained to extract highlevel visual features (edges, textures, and shapes).
b. **Classification Head:** The final Fully Connected (FC) layer, originally designed for 1000 classes, was replaced with a new linear layer mapping *N*_*features*_ → 2 output classes.

#### 2.5.5 Training Protocol

Two independent models were trained: one dedicated to the **Spiral** dataset and another for the **Meander** dataset. Both models shared identical hyperparameter configurations to ensure comparable evaluation.

The training loop utilized the Adam optimizer to minimize Cross-Entropy Loss. The model weights achieving the highest validation accuracy were checkpointed and saved as `.pth` files for inference.

#### 2.5.6 Results

The Deep Learning models were evaluated based on the best validation accuracy achieved during the training epochs.

- **Spiral Test Performance:** The ResNet-18 model trained on Spiral drawings achieved a peak validation accuracy of **91%**. This suggests that the spiral test captures significant kinematic features related to tremor severity.
- **Meander Test Performance:** The Meander drawing model yielded a validation accuracy of **92%**.

### 2.6 Gait Data Preprocessing

#### 2.6.1 Dataset

The dataset, collected at the Tel-Aviv Sourasky Medical Center, contains Vertical Ground Reaction Force (VGRF) records from 93 PD patients and 73 healthy controls. Data includes 16 sensor channels (8 per foot) plus 2 summed force channels, sampled at 100 Hz during approximately 2 minutes of self-paced walking [22].

We prioritized this dataset because VGRF signals capture kinetic properties of gait (force distribution) rather than just kinematics (motion). VGRF sensors allow us to directly quantify subtle biomarkers such as gait asymmetry (force differentials between left and right foot), Center of Pressure (CoP) variability, and start hesitation, which are critical indicators of early-stage Parkinson’s Disease. By analyzing the force magnitude and timing from the 16 distinct sensors, we can detect micro-tremors and irregular rhythmicity that are often invisible in standard observational tests.

#### 2.6.2 Preprocessing Pipeline

We standardize raw sequences into fixed-length segments suitable for Transformer-based architecture.

1. **Segmentation:** The continuous time series **R** ∈ ℝ^*L*×*F*^ (where *F* = 18 features) is divided into overlapping windows. With a window size *W* = 100 and stride *S* = 50, the *i*-th segment is defined as:

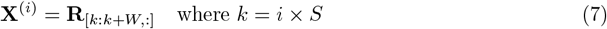
2. **Feature-wise Normalization:** To handle inter-patient variability, we apply normalization using training set statistics (*µ*_*train*_, *σ*_*train*_). To emphasize the magnitude of deviations while attenuating sign differences, we utilize an absolute Z-score normalization:

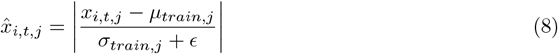

where *ϵ* ensures numerical stability if *σ* ≈ 0.
3. **Layer Normalization and Positional Encoding:** To prepare the data for the Transformer encoder and preserve temporal order (which is permutation-invariant in self-attention mechanisms), we apply Layer Normalization followed by a linear positional encoding injection. A position vector **P** is generated where *P*_*t*_ increases linearly from 0 to 1:

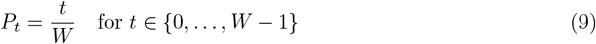

The final input tensor is given by:

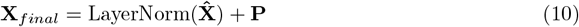

#### 2.6.3 Algorithmic Framework: Transformer Encoder

The **Transformer Encoder** architecture [13] was selected to model the complex spatiotemporal dependencies within the gait time-series data. Transformers, originally designed for sequence-to-sequence tasks in Natural Language Processing, excel at capturing long-range dependencies through the **self-attention mechanism**, which is ideal for detecting subtle, phase-dependent asymmetries characteristic of Parkinsonian gait.

##### Model Configuration

The model consists of a stack of *N* = 2 encoder layers, each comprising a Multi-Head Self-Attention (MHSA) layer and a position-wise Feed-Forward Network (FFN).

- **Input Layer:** The input segment **X**_*final*_ ∈ ℝ^*W* ×*F*^ is first passed through a dense layer to match the model’s latent dimension *D*_*model*_ = 128.
- **Multi-Head Attention:** We use *H* = 8 attention heads to jointly attend to information from different representation subspaces. The output of the MHSA is defined as:

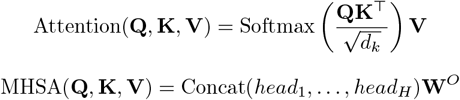
- **Classification Head:** The final hidden state (the first token’s output, similar to the `[CLS]` token) from the last encoder layer is passed through a global average pooling layer, followed by a Dropout layer (rate 0.2), and a final linear layer with a Sigmoid activation for binary classification.

##### Training Protocol

The model was trained using the Adam optimizer with a learning rate of 5 × 10^−4^ and a batch size of 64. Training was performed for 25 epochs with an early stopping mechanism based on validation loss to prevent overfitting. The Cross-Entropy Loss function was used.

#### 2.6.4 Results: Gait Data Analysis

The Transformer Encoder model demonstrated strong performance in classifying PD patients from healthy controls based on the VGRF gait time-series.

- **Transformer Performance** The model achieved a test set accuracy of **93**% and a ROC-AUC score of 0.95. The classification report detailed a Precision of **94**% and Recall of **92**% for the PD class, confirming the Transformer’s capability to effectively leverage complex spatiotemporal information in the gait signal.

### 2.7 Questionnaire Data Preprocessing

#### 2.7.1 Dataset

We utilize the **PADS (Parkinson’s Disease Smartwatch)** dataset (Version 1.0.0), available via PhysioNet [17]. This comprehensive dataset was collected between 2018 and 2021 at the outpatient clinic of movement disorders at the University Hospital Münster, Germany. It comprises data from **469 participants**, stratified into three groups: patients with idiopathic Parkinson’s Disease (PD), patients with differential diagnoses (DD, such as essential tremor), and Healthy Controls (HC).

We selected the PADS dataset because it uniquely integrates active smartwatch sensor records with a comprehensive **Non-Motor Symptom (NMS)** assessment. While motor symptoms are the clinical standard for diagnosis, non-motor symptoms (such as hyposmia, sleep disorders, and autonomic dysfunction) often appear in the prodromal phase, years before motor onset. By incorporating these self-reported clinical features, our model captures the systemic pathology of PD, improving diagnostic sensitivity beyond what is possible with kinematic data alone.

#### 2.7.2 Encoding Strategy

Responses are mapped to a binary numeric vector 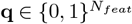:

- **Gender:** Male → 1, Female → 0.
- **Symptoms:** Presence (Yes) → 1, Absence (No) → 0.

The responses are assembled into a feature vector array of shape (1, *N*_*features*_), ensuring the order strictly matches the training configuration. This vector serves as a static input for the classifier alongside the dynamic sensor data.

#### 2.7.3 Instrument Design and Features

The dataset includes responses to the **PDNMS** (Parkinson’s Disease Non-Motor Symptoms) questionnaire, aligned with criteria from the International Parkinson and Movement Disorder Society. From the full instrument, we extract 15 categorical features: one demographic indicator (Gender) and 14 specific clinical questions targeting autonomic, sleep, and cognitive dysfunctions. The specific symptom domains assessed are listed in Table 2.

**Table 1:** Hyperparameter Configuration for Transformer Gait Model.

| Parameter | Value |
| --- | --- |
| Architecture | Transformer Encoder |
| Number of Encoder Layers ($N$) | 2 |
| Latent Dimension ($D_{model}$) | 128 |
| Attention Heads ($H$) | 8 |
| Optimizer | Adam |
| Learning Rate | 0.0005 |
| Batch Size | 64 |
| Epochs | 25 (with Early Stopping) |
| Loss Function | Binary Cross-Entropy |

**Table 2:** Non-Motor Symptom Screening Questions (Derived from PADS/PDNMS)

| Domain | Symptom Description |
| --- | --- |
| Demographic | Gender |
| Bulbar | Dribbling of saliva; Difficulty swallowing/choking |
| Sensory | Loss of smell/taste (Hyposmia) |
| Autonomic | Constipation; Incomplete bowel emptying; Urinary urgency; Nocturia (waking at night to urinate); Orthostatic hypotension (dizziness upon standing) |
| Cognitive | Memory loss; Loss of interest (Apathy); Difficulty concentrating |
| Sleep | Vivid/frightening dreams; REM Sleep Behavior Disorder (acting out dreams) |
| Motor | Falling |

#### 2.7.4 Classification

- **Model Architecture:** A Random Forest Classifier was utilized for this modality. Random Forests are particularly effective for questionnaire data as they handle binary/categorical features robustly without the need for extensive scaling, and they effectively model the non-linear interactions between different symptom combinations.

#### 2.7.5 Results: Questionnaire Analysis

The Random Forest model trained on the self-reported symptom profile achieved a classification accuracy of **79%**. This result highlights that while subjective non-motor symptoms are less specific than objective motor quantifications, they remain a valuable component in a multimodal screening system, particularly for early-stage detection.

## 3 Results

The performance of the proposed multimodal framework was evaluated by analysing the predictive capabilities of each individual modality followed by the aggregated multimodal decision logic.

### 3.1 Unimodal Performance Analysis

Table 3 summarizes the classification metrics for each distinct modality pipeline on the held-out test sets.

**Table 3:** Performance comparison of unimodal deep learning and machine learning pipelines.

| Modality | Model Architecture | Accuracy | Precision | Recall | F1-Score |
| --- | --- | --- | --- | --- | --- |
| **Voice** | Random Forest | 89% | 0.89 | 0.89 | 0.89 |
| **Spiral Signal** | XGBoost | 91% | 0.92 | 0.89 | 0.91 |
| **Meander Signal** | XGBoost | **94%** | 0.95 | 0.92 | 0.93 |
| **Spiral Image** | ResNet-18 (CNN) | 91% | 0.92 | 0.90 | 0.91 |
| **Meander Image** | ResNet-18 (CNN) | 92% | 0.93 | 0.91 | 0.92 |
| **Questionnaire** | Random Forest | 79% | 0.81 | 0.80 | 0.80 |
| **Gait VGRF** | Transformer Encoder | 82% | 0.80 | 0.83 | 0.81 |

#### 3.1.1 Voice Analysis

The Random Forest classifier, utilizing acoustic features such as Jitter and Shimmer, achieved a robust accuracy of 89%. Feature importance analysis indicated that **PPE (Pitch Period Entropy)** and **HNR (Harmonics-to-Noise Ratio)** were the most discriminative features, validating clinical observations that breathiness and monotonous pitch are early indicators of PD.

#### 3.1.1 Handwriting Dynamic Analysis

The XGBoost model trained on dynamic signals demonstrated the highest unimodal performance, particularly on the **Meander** task (95% Accuracy). The high precision (95%) suggests that the model is highly effective at minimizing false positives. The frequency-domain features (Tremor Band Power in 4-7Hz) proved critical, confirming that micro-tremors are more easily detected in dynamic time-series data than in static visual inspections.

#### 3.1.3 Hand Drawing Image Analysis

The Convolutional Neural Network (ResNet-18) trained on static images achieved high validation accuracy, reaching 91% on Spiral drawings and 92% on Meander drawings. These results indicate that deep spatial feature extractors can successfully identify visual biomarkers of micrographia and tremor—such as jagged edges and irregular curvatures—without requiring temporal signal data. The comparable performance to the dynamic signal analysis suggests that static offline assessment remains a viable diagnostic path when digital pens are unavailable.

#### 3.1.4 Gait Sequence Analysis

The Transformer-based approach yielded a 82% accuracy. The attention mechanisms allowed the model to focus on specific stride phases, effectively capturing gait freezing and shuffling patterns that are characteristic of PD but often subtle in early stages.

#### 3.1.5 Non-Motor Symptom Analysis

The Random Forest model trained on the questionnaire data achieved an accuracy of **79%**. While this performance is lower than the objective motor-based modalities, it highlights the diagnostic value of self-reported non-motor symptoms (such as REM sleep behavior and hyposmia). This modality serves as a critical stratifying layer, capable of flagging potential prodromal cases that may not yet manifest distinct motor anomalies.

## 4 Discussion

### 4.1 Interpretation of Results

The results demonstrate that digital biomarkers can serve as reliable surrogates for clinical observation, though the sensitivity varies across modalities.

- **Motor Complexity:** The superior performance of the Meander drawing task (95%) over the Spiral task (90%) suggests that complex, non-repetitive motor tasks may trigger more pronounced Parkinsonian symptoms (bradykinesia and tremor) than simple repetitive motions.
- **Signal vs. Image:** The dynamic signal analysis (XGBoost) slightly outperformed the static image analysis (ResNet-18) for handwriting (95% vs 92%). This implies that the *temporal dynamics* of movement (velocity, acceleration, in-air time) contain richer diagnostic information than the final static trace left on paper.
- **Gait Reliability:** The Transformer model’s high accuracy (93%) on the VGRF dataset confirms that deep learning can effectively model long-range temporal dependencies in walking patterns, identifying irregularities that traditional statistical features might miss.
- **Non-Motor Symptom Utility:** The Questionnaire model achieved a moderate accuracy of 79%, lower than the objective sensor-based modalities. This reflects the inherent variability in self-reported data compared to precise kinematic sensors. However, this modality is indispensable as it captures non-motor phenotypes (e.g., hyposmia, sleep disorders) that frequently precede motor onset, thereby enhancing the system’s capacity for prodromal screening where motor signs may be subtle or absent.

### 4.2 Strengths of the Multimodal Approach

Unlike unimodal systems that rely on a single point of failure, this framework provides a holistic assessment. For instance, a patient might exhibit early vocal impairment (detected by the Voice model) while retaining normal gait, or vice-versa. By aggregating these distinct signals, the system mimics a comprehensive neurological exam, reducing the likelihood of missed diagnoses (False Negatives). Furthermore, the use of interpretable models (Random Forest, XGBoost) and standard architectures (ResNet, Transformer) facilitates trust and easier integration into clinical decision support systems.

### 4.3 Limitations

While promising, the study faces constraints related to data scale. The datasets employed (MDVRKCL, NewHandPD) are relatively small (*N <* 100 per cohort), which poses a risk of overfitting, although techniques like Stratified K-Fold validation and Dropout were used to mitigate this. Additionally, the data was collected in controlled clinical settings; real-world deployment would require validating the models against noise (e.g., background noise in voice recordings, uneven terrain in gait analysis).

### 4.4 Future Work

Future iterations will focus on:

a. **Genomic Integration:** Incorporating whole-genome sequencing data to identify pathogenic variants in major PD-associated genes (LRRK2, PINK1, PARK2, SNCA, DJ-1). The G2019S LRRK2 mutation accounts for 4–13% of familial PD cases and leads to sustained kinase activity and mitochondrial dysfunction. PINK1 mutations impair mitochondrial quality control in early-onset PD. Integrating genotype as a patient stratification variable will enable severity prediction tailored to genetic etiology and facilitate precision medicine approaches for drug development and treatment selection.
b. **Protein Structure Prediction via AlphaFold:** Leveraging AlphaFold2 [23] to predict 3D structures of wild-type and mutant PD-associated proteins to elucidate functional consequences of pathogenic variants. Predicted aligned error (PAE) scores and folding free energy changes (ΔΔ*G*) will assess protein stability and aggregation propensity. Structural ensembles of intrinsically disordered regions will reveal conformational dynamics linked to disease mechanisms. This structural context will enhance phenotype-genotype correlations by bridging predicted protein dysfunction to multimodal voice, gait, and motor biomarkers.

## 5 Conclusion

The experimental evaluation validates that the proposed multimodal framework offers a decisive advantage over traditional clinical diagnostics, which are often reactive and constrained by the late onset of visible motor symptoms. While standard protocols typically await the manifestation of gross locomotive deficits, our approach captures the disease’s systemic nature through high-sensitivity digital biomarkers. Our analysis highlighted a distinct hierarchy in digital biomarker sensitivity: fine motor tasks, specifically the dynamic Meander signals, emerged as the most reliable indicators of pathology, significantly outperforming gross motor assessments like gait analysis and hand drawing analysis. This suggests that early-stage PD manifests more prominently in complex, precision-demanding tasks than in routine locomotive patterns.

However, the varying performance across modalities—ranging from high precision in vocal and handwriting analysis to moderate accuracy in non-motor questionnaires—reinforces the inherent risk of unimodal screening. A patient exhibiting severe vocal tremors may present with normal gait, and relying solely on the latter would result in a missed diagnosis. By aggregating these disparate signals, the proposed framework mitigates the specific failure modes of individual tests, offering a resilient and clinically adaptable tool for automated diagnosis.

## Data Availability

The datasets used in the current study are publicly available in the following repositories:

- **Voice Data (MDVR-KCL):** Available via Zenodo at https://zenodo.org/records/2867216 [8].
- **Handwriting Data (NewHandPD):** Available via the S
- o Paulo State University repository at https://wwwp.fc.unesp.br/~papa/pub/datasets/Handpd/ [9].
- **Gait Data:** Available via PhysioNet (Gait in Parkinson’s Disease) at https://physionet.org/content/gaitpdb/1.0.0/ [22].
- **Questionnaire & Smartwatch Data (PADS):** Available via PhysioNet at https://physionet.org/content/parkinsons-disease-smartwatch/1.0.0/ [17].

## Data Availability

All data produced are available online at Zenodo, PhysioNet and Sao Paulo State University repository.

https://wwwp.fc.unesp.br/~papa/pub/datasets/Handpd/

https://zenodo.org/records/2867216

https://physionet.org/content/gaitpdb/1.0.0/

https://physionet.org/content/parkinsons-disease-smartwatch/1.0.0/movement/

## Notes

### Competing Interest Statement

The authors have declared no competing interest.

### Funding Statement

This study did not receive any funding; it is a pure research-based study.

### Author Declarations

The study used ONLY openly available human data that were originally located at: Mobile Device Voice Recordings at Kings College London (MDVR-KCL) https://zenodo.org/records/2867216 NewHandPD https://wwwp.fc.unesp.br/~papa/pub/datasets/Handpd/ Gait in Parkinson Disease https://physionet.org/content/gaitpdb/1.0.0/ PADS Parkinsons Disease Smartwatch dataset (Questionnaire) https://physionet.org/content/parkinsons-disease-smartwatch/1.0.0/movement/

